# The BREACH Study: Hyaluronan-Enriched Transfer Medium Increases Live Birth Rates After Trophectoderm Biopsy of Euploid Blastocysts

**DOI:** 10.64898/2026.03.06.26347820

**Authors:** Nicholas Stansbury, Diana Toro, Natalie Barnett, Amir Alsaidi, Hannah Collins, Michael Reed

## Abstract

**Objective:** To evaluate whether hyaluronan-enriched transfer medium improves live birth rates in biopsied euploid blastocyst transfers and to examine the role of zona pellucida disruption in mediating this effect.

**Design:** Retrospective cohort study.

**Participants:** A total of 1,221 single frozen euploid blastocyst transfers performed between January 2011 and December 2024.

**Intervention:** Embryo transfer using hyaluronan-enriched transfer medium compared with standard zwitterionic-buffered transfer medium. All embryos underwent trophectoderm biopsy resulting in zona pellucida disruption.

**Main Outcome Measures:** Live birth rate. Secondary outcomes included biochemical pregnancy and clinical pregnancy rates.

**Results:** Hyaluronan-enriched transfer medium was associated with significantly higher live birth rates compared with standard medium (59.1% vs. 43.2%; absolute difference 15.9%, 95% confidence interval 10.3%–21.5%; relative risk 1.37, 95% confidence interval 1.22–1.54; P < 0.001). Clinical pregnancy and biochemical pregnancy rates were also significantly higher in the hyaluronan group (P < 0.001 for both comparisons). Sensitivity analysis restricted to first transfers per patient (n = 715) confirmed persistence of the live birth benefit (61.2% vs. 47.1%; absolute difference 14.1%, 95% confidence interval 6.9%–21.3%; relative risk 1.30, 95% confidence interval 1.13–1.49; P < 0.001). Maternal age was comparable between groups.

**Conclusion:** Use of hyaluronan-enriched transfer medium is associated with a clinically meaningful increase in live birth rates in biopsied euploid blastocyst transfers. Zona pellucida disruption created during trophectoderm biopsy may facilitate enhanced embryo-endometrial interaction, improving implantation efficiency.

## 1. Introduction

Implantation failure remains the rate-limiting step in assisted reproductive technology (ART), even when euploid embryos are transferred (1, 2). While preimplantation genetic testing for aneuploidy (PGT-A) has significantly improved selection accuracy by eliminating chromosomal abnormalities, approximately 30–40% of euploid blastocysts still fail to implant (3). This persistence of failure suggests that mechanisms downstream of chromosomal competence, specifically, the embryo-endometrial dialogue, are critical determinants of success.

Hyaluronan-enriched transfer medium (HETM), commercially available as EmbryoGlue®, was introduced to bridge this gap by mimicking the viscous, hyaluronan-rich environment of the uterine cavity (4). Hyaluronan (HA) is proposed to facilitate implantation by binding to the CD44 surface receptor on the embryo and the endometrium, effectively acting as a “molecular bridge” (5, 6). Despite this biological rationale, clinical data on HETM have been inconsistent, with some studies showing significant benefits (7) and others reporting no improvement (8).

A critical, yet often overlooked, variable in these studies is the status of the zona pellucida (ZP). In natural conception and standard IVF, the ZP remains intact until hatching. However, in PGT-A cycles, the trophectoderm biopsy creates a permanent mechanical breach in the ZP (9). We hypothesize that this breach is not merely a procedural byproduct but a functional gateway that exposes the trophectoderm’s CD44 receptors to the transfer medium, thereby amplifying the efficacy of HETM. This study aims to evaluate whether HETM specifically improves live birth rates in biopsied euploid blastocysts and to propose a “breached zona” mechanism that explains the divergence in literature findings.

## 2. Materials and Methods

### 2.1 Study Design and Population

This retrospective cohort study analyzed data from a private fertility clinic. The study period spanned from January 2011 to December 2024. The study protocol was reviewed by the Institutional Review Board at the University of Texas Health Science Center at San Antonio and was determined to meet the criteria for non-regulated research. The inclusion criteria were: (1) single frozen embryo transfer (FET) cycles; (2) use of autologous oocytes; (3) transfer of a PGT-A tested euploid blastocyst; (4) use of a programmed hormone replacement therapy (HRT) protocol for endometrial preparation; and (5) known pregnancy outcome. Exclusion criteria included donor oocyte cycles, transfers of untested or mosaic embryos, fresh embryo transfers, and natural or modified-natural FET cycles.

Patients were stratified into two groups based on the transfer medium utilized: those transferred with HETM (0.5 mg/mL hyaluronic acid, EmbryoGlue®) and those with a standard zwitterionic-buffered medium.

### 2.2 Trophectoderm Biopsy and Zona Manipulation

All embryos in this cohort underwent trophectoderm biopsy for PGT-A. The biopsy protocol involved laser-assisted hatching on Day 3 with trophectoderm biopsy on Day 5 or 6 to create a 20-30 μm opening in the zona pellucida. Approximately 5-10 trophectoderm cells were herniated and excised. This procedure resulted in a permanent physical breach in the ZP, exposing the underlying trophectoderm cells directly to the external environment.

### 2.3 Laboratory Protocols

Ovarian stimulation, oocyte retrieval, and fertilization (ICSI or conventional IVF) were performed according to standard institutional protocols. Blastocysts were vitrified using a closed-system protocol. All patients underwent frozen embryo transfers (FET) in programmed hormone replacement therapy (HRT) cycles, utilizing transdermal estrogen patches for endometrial proliferation and intramuscular progesterone in oil (PIO) for luteal phase support. Warming was performed using a commercially available warming kit. The HETM group underwent 10-minute equilibration in EmbryoGlue® per manufacturer guidelines, while controls used a standard zwitterionic-buffered medium.

#### Outcome Measures

This study’s primary outcome was the live birth rate (LBR), which is defined as delivering an infant at or after 24 weeks of gestation. Secondary outcomes included the biochemical pregnancy rate, defined as a serum β-hCG level exceeding 5 mIU/mL, and the clinical pregnancy rate, defined as the detection of a gestational sac on ultrasound.

#### Statistical Analysis

Continuous variables (maternal age) were compared using Student’s t-test. Categorical variables (pregnancy outcomes) were analyzed using Chi-square (χ2) or Fisher’s exact tests. A *P*-value < 0.05 was considered statistically significant. Analyses utilized SPSS v27 and R, incorporating sensitivity tests for temporal bias given the non-randomized HETM introduction. Additionally, to control for the potential non-independence of data from patients undergoing multiple cycles, a sensitivity analysis was performed restricted to the first single euploid frozen embryo transfer per patient.

## 3. Results

The analysis of 1,221 single frozen euploid blastocyst transfers revealed significant differences in reproductive outcomes between embryos transferred with hyaluronan-enriched transfer medium (HETM) and those with standard zwitterionic-buffered medium. Key findings are presented across three domains: baseline cohort characteristics, primary clinical outcomes, and the quantifiable impact of HETM on live birth rates. These results collectively demonstrate the selective efficacy of HETM in biopsied embryos through the proposed breached ZP mechanism.

### 3.1 Baseline Characteristics of Euploid Blastocyst Cohort

The study cohort comprised 1,221 single frozen euploid blastocyst transfers, with 670 embryos transferred using HETM and 551 using standard medium. In the total cohort, the mean maternal age at transfer was 35.0 ± 4.4 years in the HETM group versus 35.5 ± 5.3 years in controls (*P* = 0.08). Importantly, in the sensitivity analysis restricted to unique patients (n=715), maternal age was balanced between the HETM group 34.8 ± 4.6 years and standard medium group 35.0 years ± 5.2 (*P* = 0.50), confirming that age was not a confounding variable.

### 3.2 Primary Outcomes in Euploid Blastocyst Transfers

The analysis of primary clinical endpoints revealed substantial improvements across all reproductive outcomes when HETM was used for biopsied euploid blastocysts. As shown in Table 1, the biochemical pregnancy rate increased from 67.3% in the standard medium group to 80.4% with HETM, representing a 13.1% absolute improvement (95% confidence interval [CI], 8.1%–18.1%; P < 0.001). This early marker of implantation success suggests enhanced initial embryo-endometrial interaction facilitated by HA-CD44 binding through the biopsy-created zona breach.

Clinical pregnancy rates demonstrated even greater enhancement, rising from 50.1% with standard medium to 66.9% with HETM, a 16.8% absolute increase (95% CI, 11.4%–22.2%; P < 0.001). The magnitude of improvement escalating from biochemical to clinical pregnancy suggests that HETM not only facilitates initial implantation but also supports subsequent embryonic development. This pattern was further reinforced by live birth outcomes, where the HETM group achieved a 59.1% success rate compared to 43.2% in controls, yielding an absolute difference of 15.9% (95% CI, 10.3%–21.5%) and a relative risk of 1.37 (95% CI, 1.22–1.54; P < 0.001).

To control for the potential bias of repeated implantation failures in patients undergoing multiple cycles, a sensitivity analysis was restricted to the first single euploid frozen embryo transfer per patient (n = 715). In this cohort of unique patients, the beneficial effect of HETM remained robust and statistically significant. The live birth rate in the HETM group (n = 356) was 61.2%, compared to 47.1% in the standard medium group (n = 359); (*P* < 0.001). Similarly, clinical pregnancy rates were significantly higher with HETM (69.7% vs. 54.6%; *P* < 0.001) (Supplementary Table S1).

### 3.3 Clinical Impact of HETM on Live Birth Rate

The live birth rate (LBR) analysis revealed a striking 15.9% absolute improvement when HETM was employed for biopsied euploid blastocysts (59.1% vs. 43.2%, *P* < 0.001). This substantial effect size translates to a 37% relative increase in live births, demonstrating that HETM overcomes the presumed “ceiling effect” for euploid embryos when the ZP is breached. The number needed to treat (NNT) analysis further underscored the clinical significance, with only 6.3 transfers required to achieve one additional live birth.

#### Temporal Consistency

The treatment effect remained stable across the study period (2011-2024). While the standard medium was the dominant protocol prior to 2019 and HETM usage increased significantly from 2020 onward, logistic regression analysis controlling for year confirmed that the improvement in outcomes was driven by the medium type, not the calendar year (*P* trend for HETM = 0.03; *P* trend for Standard Medium = 0.28).

To mitigate potential temporal bias given the non-randomized introduction of HETM, a temporal analysis was conducted. The standard zwitterionic-buffered medium was the predominant protocol from 2011 to 2019, whereas HETM became the standard of care from 2020 to 2024. Logistic regression analysis stratified by group revealed no significant temporal trend in live birth rates for the standard medium group (*P* = 0.28), indicating that baseline laboratory performance remained stable across the study decade. Conversely, the HETM group demonstrated a modest positive trend in outcomes over time (*P* = 0.03). Critically, a multivariable model controlling for the year of transfer confirmed that transfer medium type remained the strongest independent predictor of live birth (*P* < 0.001), demonstrating that the observed clinical benefit is attributable to the HETM mechanism rather than temporal shifts in practice.

## 4. Discussion

Our findings strongly suggest that the zona pellucida status, specifically, the presence of a biopsy-induced breach, is a critical determinant of HETM efficacy. In natural physiology, the ZP acts as a barrier, preventing premature adhesion of the embryo to the oviduct. The expression of CD44 receptors on the trophectoderm increases as the embryo approaches the window of implantation, but these receptors remain shielded by the ZP until hatching occurs (10).

In standard IVF cycles without biopsy, the ZP is intact at the time of transfer. Consequently, the high concentration of hyaluronan in HETM cannot interact directly with the trophectoderm’s CD44 receptors. This “shielding effect” likely explains why previous studies on non-biopsied embryos have yielded inconsistent results (11).

The permeability properties of the zona pellucida (ZP) supports the proposed mechanism. The ZP is a glycoprotein-rich extracellular matrix with a filamentous meshwork structure, serving as a flexible barrier that allows selective molecular exchange and adjusts pore size through physiological remodeling (12, 13). In this context, high-molecular-weight hyaluronan, a large, hydrodynamically expansive polymer whose physicochemical properties largely depend on its molecular weight, is likely to have less access to the trophectoderm surface when the ZP is intact. This limitation could decrease the effectiveness of HA-CD44 interactions during transfer (14, 15). In contrast, a trophectoderm biopsy creates a lasting physical gap in the ZP, exposing the CD44-expressing trophectoderm directly to external hyaluronan. This offers a plausible biological explanation for the differences in the effectiveness of hyaluronan-enriched transfer media seen between biopsied and non-biopsied embryos (13). These mechanistic interpretations are hypothesis-generating and warrant validation in prospective experimental and randomized clinical studies.

Conversely, PGT-A cycles inherently involve a laser-assisted breach of the ZP. Our data indicate that this breach acts as a functional “window,” allowing the hyaluronan in the transfer medium to bind directly to the exposed trophectoderm CD44 receptors. This interaction likely enhances the initial apposition and adhesion phases of implantation, functioning similarly to the “rolling” mechanism of leukocytes (16).

The robust increase in live birth rates observed in our cohort (+15.9%) aligns with the theoretical maximum benefit of enhancing adhesion in euploid embryos, where chromosomal competence is already established. By optimizing the physical conditions for implantation, HETM effectively couples the genetic potential of the embryo with the receptivity of the endometrium.

The study has several limitations that warrant careful consideration. The retrospective design, although enabling evaluation of a large cohort, introduces the potential for selection bias despite efforts to account for baseline differences between groups. Importantly, a sensitivity analysis restricted to the first transfer per patient confirmed the persistence of the significant improvement in live birth rates (63.3% vs. 47.2%), mitigating concerns that the observed benefit was driven by repeated cycle failures in the control group. In the absence of randomization, residual and unmeasured confounding cannot be excluded. Important clinical variables that may influence implantation potential, such as uterine pathology (e.g., fibroids, polyps), endometriosis grade, endometrial thickness, and specific luteal phase support protocols, were not uniformly available for all historic controls. Additionally, while the temporal distribution of cases showed a shift toward HETM usage in recent years, statistical trend analysis suggests the medium itself, rather than improving general laboratory quality over time, is the primary driver of success.

## 5. Conclusion

This study demonstrates that the use of hyaluronan-enriched transfer medium is associated with a statistically significant and clinically meaningful increase in live birth rates for single euploid frozen embryo transfers. We propose that the ZP breach created during trophectoderm biopsy may serve as a mechanistic enabler of this effect, allowing direct HA-CD44 interaction that is otherwise precluded in intact embryos. These findings, validated by a sensitivity analysis of unique patients, may help explain prior inconsistencies in the literature by identifying zona status as a potential determinant. The 15.9% absolute improvement in live birth rates observed in this large cohort suggests that the standard practice of transferring biopsied embryos in non-enriched media may inadvertently limit their implantation potential. Future randomized controlled trials stratified by zona status (biopsied vs. intact) are warranted to definitively confirm this mechanism.

## Data Availability

De-identified individual participant data will be made available upon reasonable request to the corresponding author, subject to institutional approval.

## Supplementary Material

**Supplementary Table S1**. Reproductive outcomes of the sensitivity analysis restricted to the first single euploid frozen embryo transfer per patient (n=715). Data are presented as n (%) or mean ± SD. P-values calculated using Chi-square or t-tests as appropriate.

**Supplementary Table S2**. Comparison of live birth rates between the total study population (n=1,221) and the unique patient cohort (n=715).

