## Supplementary Tables S1-S2 for "The BREACH Study: Hyaluronan-Enriched Transfer Medium Increases Live Birth Rates After Trophectoderm Biopsy of Euploid Blastocysts"

**Supplementary Data:**

**Supplementary Table S1**

**Table 1. Live Birth Rate by Transfer Medium**

| **Analysis Population** | **HETM Live Birth Rate** | **Standard Medium Live Birth Rate** | **Absolute Difference** | **P-value** |
| --- | --- | --- | --- | --- |
| All Transfers (n = 1,221) | 59.1% (396/670) | 43.2% (238/551) | +15.9% | < 0.001 |
| First Transfer Only (n = 715) | 61.2% (218/356) | 47.1% (174/359) | +16.1% | < 0.001 |

**Note.** The “First Transfer Only” analysis excludes repeat cycles from the same patient to ensure statistical independence.

**Table 2. Clinical Outcomes – First Transfer Only Cohort**

| **Variable** | **HETM Group (n = 356)** | **Standard Medium Group (n = 359)** | **P-value** |
| --- | --- | --- | --- |
| Maternal age (years) | 34.8 ± 4.6 | 35.0 ± 5.2 | 0.50 |
| Biochemical pregnancy | 299/356 (84.0%) | 256/359 (71.3%) | < 0.001 |
| Clinical pregnancy | 248/356 (69.7%) | 196/359 (54.6%) | < 0.001 |
| Live birth rate | 218/356 (61.2%) | 169/359 (47.1%) | < 0.001 |
